# LGBTQ+ Affirming Care May Increase Awareness and Understanding of Undetectable=Untransmittable among Midlife and Older Gay and Bisexual Men in the US South

**DOI:** 10.1101/2022.06.20.22276658

**Authors:** Tara McKay, Ellesse-Roselee Akré, Jeff Henne, Adam Conway, Isabel Gothelf, Nitya Kari

## Abstract

One of the most significant innovations in HIV prevention is the use of HIV treatment to prevent HIV transmission. This information has been disseminated as the “Undetectable = Untransmittable” (U=U) message. Despite evidence of effectiveness, U=U awareness, belief, and understanding remains limited in some communities. In this study we examine whether having an LGBTQ affirming healthcare provider increases U=U awareness, belief, and understanding among midlife and older gay and bisexual men in the US South, an understudied and underserved population and region where new HIV infections are increasing. We use data from the Vanderbilt University Social Networks Aging and Policy Study (VUSNAPS) on sexual minority men aged 50 to 76 from four Southern US states collected in 2020-2021. We find that only one in four men report prior awareness of U=U, but awareness is higher among HIV negative and HIV positive men who have an LGBTQ affirming provider. Having an affirming provider significantly increases U=U belief and understanding, improves risk perception accuracy, and increases the likelihood of having ever tested for HIV among HIV negative men. Improving access to LGBTQ affirming healthcare may improve U=U awareness, belief, and understanding, which could help to curb HIV transmission in the US South.

---

One of the most significant innovations in HIV prevention in the last two decades has been the use of HIV treatment to prevent HIV transmission. The resulting global public health campaign, “Undetectable equals Untransmittable”, or U=U, underscores the importance of achieving and maintaining viral suppression in people living with HIV to prevent HIV transmission. By taking antiretroviral therapy (ART) daily as prescribed, people with HIV cannot sexually transmit the virus to others [1]. In the US, this campaign has been widely supported by the Centers for Disease Control and Prevention, the National Institute of Allergy and Infectious Diseases, and the American Medical Association [1–3].

Large-scale survey studies using data collected from community-based samples of people living with HIV and men who have sex with men generally find very high levels of awareness of the U=U concept in the US [4,5] and observe increases in awareness of U=U over the last decade, especially among HIV positive men [5,6]. Among HIV negative men surveyed from 2017 and 2018 in the US, 85% report being aware of U=U [7].

However, several studies of men who have sex with men in the US suggest that understanding and application of U=U are substantially more limited. Online surveys of men who have sex with men consistently find that just three to four out of every ten men who have sex with men correctly identify HIV treatment or viral suppression as providing protection against transmission [8–10].

Importantly, existing studies have not generally focused on U=U awareness and understanding among midlife and older sexual minority men. The median age of most U=U awareness studies sampling sexual minority men in the US is consistently younger than 40 and, for some, younger than 35 or even 30 [4,7,9,10]. One study that explicitly assessed age-cohort differences in HIV prevention knowledge, risk perception, and behaviors among gay and bisexual men in the US found that men in younger age-cohorts had greater functional knowledge of HIV prevention strategies, including condom use, pre-exposure prophylaxis (PrEP), post-exposure prophylaxis (PEP), and Treatment as Prevention/U=U [8].

Additionally, among studies with adequate sample sizes to test geographic variation within the US, HIV negative men living in Southern states in the US were significantly less likely to have heard of U=U [7]. In another study, HIV positive men in Southern states were less likely to rate the U=U concept as accurate compared to HIV positive men in the Northeast and Western states [11]. These gaps are important because Southern states comprise more than 50% of new HIV infections, most of which are among men who have sex with men [12]. Additionally, fewer people in the South are aware that they have HIV compared to other US regions, delaying access to treatment, and there has been lower uptake of other medical prevention technologies, like PrEP [12,13].

In this study we address gaps in the U=U landscape and expand on existing work by examining healthcare-related determinants of U=U awareness, understanding, and impact among midlife and older sexual minority men in the US South. Although many sexual minority men hear about the U=U message from sources other than a healthcare provider, healthcare providers are an important point of regular contact for HIV-positive men on treatment and HIV-negative men seeking sexual healthcare services or who are taking pre-exposure prophylaxis (PrEP). However, a substantial share of sexual minority men do not talk to their healthcare providers about having sex with men and, thus, may not be receiving adequate or appropriate sexual health care and information [14,15]. A recent national study found that, among men who have sex with men in the US, 30% of those with a primary care provider reported that they had not disclosed their sexual orientation to their primary healthcare provider [7].

Patients may not disclose their sexual behavior or identity for several reasons, including because providers do not ask, past negative experiences, fear of homophobia and stigmatization, internalized stigma, and belief that health is not related to their identity [16,17]. In a study of the Veterans Health Care Administration (VHA), more than one-third of gay, lesbian, and bisexual veterans (36.9%) reported that VHA staff “definitely does not know” about their sexual orientation and a quarter (25.1%) reported avoiding seeking services because of concerns about confidentiality, stigma, or acceptance of their sexual orientation [18].

There are also challenges to providing LGBTQ affirming care on the provider side. Today, the American Medical Association openly advocates for inclusion and nondiscrimination of LGBTQ+ patients and providers [20]. Although acceptance of LGBTQ people varies by physician specialty [21] and by reported versus implicitly held beliefs [22], studies generally find that many providers’ attitudes towards LGBTQ people are positive and have improved over time [23,24]. Nonetheless, many physicians still have difficulty providing affirming care because they were not trained to do so. The median number of hours dedicated to LGBTQ topics in medical schools is 5 hours and is largely decided by the individual institution [25,26]. As a result, many practitioners find it difficult to use unfamiliar sexual and gender terms, decide which ward to nurse transgender patients, discuss interpersonal violence and abuse, and identify LGBTQ health care resources despite otherwise holding positive attitudes toward LGBTQ people [16,27].

A lack of fluency in LGBTQ health, identities, and behaviors among providers can lead patients to delay or forgo care, even care that is not related to their LGBTQ identity or sexual health, and to not disclose their LGBTQ identities to providers [16,17,28]. Older sexual minority adults are particularly likely to report personal experiences or expectations of discrimination in healthcare settings, leading to delays in accessing care or forgone care [19]. For providers, patient nondisclosure or lack of comfort discussing sexual health issues can lead to the provision of inappropriate care, inattention to specific health care needs, missed diagnostic screenings, and less focus overall on creating LGBTQ inclusive healthcare environments in entire practices [29,30]. Specifically for sexual health, sexual minority men who do not disclose their sexual orientation to their primary care provider are less likely to have been tested for HIV in the previous two years, less likely to have been tested for gonorrhea and syphilis, and less likely to have been vaccinated against hepatitis A and B [15,31,32]. The lack of affirming care options for sexual minorities can also lead to healthcare fragmentation. Gay and bisexual men often seek care from providers outside of primary care contexts, especially for their sexual health needs, because of gaps in provider knowledge, greater comfort with community providers, financial cost, or expectations of discrimination [33].

The barriers to accessing and providing LGBTQ affirming care may be particularly acute in Southern US states. Southern states are more likely than Northeastern and Western states to have laws that explicitly exclude or do not provide adequate care for sexual and gender minorities in healthcare [34]. Southern states also have fewer “LGBTQ Healthcare Equality Leaders” compared to Northeast and Western states, according to the Human Rights Campaign 2020 Healthcare Equality Index [35]. LGBTQ affirming healthcare providers are more likely to have explicit employee and patient nondiscrimination policies as well as staff training in LGBTQ patient-centered care [35]. Lack of nondiscrimination policies perpetuates discriminatory behaviors such as verbal abuse and refusal to provide care, which deter patients and limit them from obtaining essential care [16].

Based on this prior work, we expect that sexual minority men with LGBTQ affirming providers may have different outcomes in relation to key sexual health and HIV prevention information. We test this hypothesis using original data on U=U awareness, understanding, and belief among HIV negative and HIV positive midlife and older gay and bisexual men in four Southern states in the US.

## Data and Methods

To examine the relationship among use of LGBTQ affirming care and knowledge and support of U=U among gay and bisexual men in the US South, we use data from the Vanderbilt University Social Networks, Aging, and Policy Study (VUSNAPS). VUSNAPS is a panel study of 1,256 older LGBTQ adults aged 50 to 76 residing in Alabama, Georgia, North Carolina, Tennessee. Participants were recruited using purposeful online and venue-based sampling, linked referral, and community outreach to organizations serving LGBTQ, men who have sex with men, and older populations in each state. Wave 1 was fielded from April 1, 2020 to September 30, 2021. In this study, we restrict analyses to VUSNAPS respondents who identify as gay or bisexual and who identify as men who were born male (N=676).

### U=U Measures

The VUSNAPS survey instrument includes several items to gauge awareness, belief, application, and impact of the U=U message.

We measure *awareness* using the item “Have you heard about U=U?” (1=yes, 0=no/don’t know). We also ask respondents who indicated having heard of U=U to identify where they heard of the concept.

We measure *belief* in U=U using a detailed item that explains the U=U concept. Respondents were then asked to “to rate how much you believe the U=U concept” on a Likert scale from 1 “Very unbelievable” to 5 “Very believable”. This item was recoded to 1=“somewhat believable” or “very believable” versus all others=0.

To measure understanding of U=U participants read the following vignette: “Please imagine a situation in which an HIV-positive man, who is taking medications and reduced his viral load to a point where it is undetectable, has unprotected anal sex with an HIV-negative man. The HIV-positive man is the top and he ejaculates inside the HIV-negative man. How likely do you think it is that the HIV-negative man will get the HIV virus from this encounter?” Response categories included a 7-point Likert scale from 1 “No chance or almost no chance”, 2 “Very unlikely”, 3 “Somewhat unlikely”, 4 “Not sure”, 5 “Somewhat likely”, 6 “Very likely”, 7 “Certain or almost certain”. For logistic analyses we collapse responses “No chance” and “very unlikely=1 (correct understanding) versus all others=0.

Finally, we measure *impact* of U=U on risk perception using the item “I would feel safe having sex with someone who is HIV-positive as long as he is receiving treatment and has reduced his viral load to a point where it is undetectable.” Respondents rated their level of agreement on a 5-point Likert scale from “Strongly disagree” to 5 “Strongly agree”. For logistic analyses we collapse “Agree” and “Strongly agree”=1 versus all others=0.

### Affirming Care Measures

The VUSNAPS instrument includes several measures to assess healthcare access and utilization, including whether the respondent has a primary provider, whether the respondent’s primary care provider is LGBTQ affirming, use of a secondary healthcare provider that is LGBTQ+ affirming, and reasons for not having an LGBTQ affirming provider if no affirming provider is identified.

### Covariates

We include demographic characteristics, including age, education (high school or less, some college, college degree, and graduate or professional degree), and race/ethnicity (white, black, other person of color), HIV status, and, for HIV negative men, whether they have ever had an HIV test.

### Analytic Strategy

We conduct descriptive analyses and logistic regression analyses stratified by HIV status for each U=U outcome identified above controlling for state of residence, age, education, race/ethnicity, whether the respondent has a partner or spouse, whether the respondent reported hearing of U=U prior to the survey, and whether they have ever tested for HIV (HIV negative men only).

## Results

Table 1 presents the distribution of the sample across demographic and geographic characteristics for HIV negative and HIV positive sexual minority men in the VUSNAPS sample.

**Table 1.** Characteristics of Older HIV Positive and HIV Negative Gay and Bisexual Men in the US South, VUSNAPS

|  | Overall<br>(N=676) |  | HIV Negative<br>(N=538) |  | HIV Positive<br>(N=138) |  |
| --- | --- | --- | --- | --- | --- | --- |
|  | N | % | N | % | N | % |
| Has LGBTQ Affirming Provider | 422 | 64.8 | 305 | 58.99 | 117 | 87.31 |
| Ever Tested for HIV | 556 | 82.3 | 441 | 81.97 | -- |  |
| State of Residence |  |  |  |  |  |  |
| Alabama | 119 | 17.6 | 93 | 17.29 | 26 | 18.84 |
| Georgia | 177 | 26.2 | 132 | 24.54 | 45 | 32.61 |
| North Carolina | 182 | 26.9 | 154 | 28.62 | 28 | 20.29 |
| Tennessee | 198 | 29.3 | 159 | 29.55 | 39 | 28.26 |
| Age (M/sd) | 59.3 | 6.30 | 59.6 | 6.38 | 58.5 | 5.98 |
| Education |  |  |  |  |  |  |
| High School or less | 31 | 4.7 | 20 | 3.83 | 11 | 8.21 |
| Some College | 154 | 23.5 | 113 | 21.65 | 41 | 30.60 |
| College Degree | 224 | 34.2 | 179 | 34.29 | 45 | 33.58 |
| Graduate/Professional Degree | 247 | 37.7 | 210 | 40.23 | 37 | 27.61 |
| Race/Ethnicity |  |  |  |  |  |  |
| White | 592 | 87.6 | 488 | 90.71 | 104 | 75.36 |
| Black | 47 | 7.0 | 27 | 5.02 | 20 | 14.49 |
| Other Person of Color | 37 | 5.5 | 23 | 4.28 | 14 | 10.14 |
| Partnered | 401 | 59.3 | 346 | 64.31 | 55 | 39.86 |

### LGBTQ Affirming Care

About two of every three men in the study (64.8%) identified a primary or secondary healthcare provider as LGBTQ affirming. Just over half of HIV negative men (59.0%) identified their healthcare provider as LGBTQ affirming compared with almost all (87.3%) of HIV positive men. After adjusting for state of residence and demographic characteristics (age, education, race/ethnicity), HIV positive men were more than 7 times more likely to identify their healthcare provider as LGBTQ affirming compared with HIV negative men (OR=7.10; 95% CI=3.94-12.80; see Table 2). Having an LGBTQ affirming care provider increased the odds that HIV negative men reported ever testing for HIV by more than 2 times (OR=2.26; 95% CI=1.38-3.72).

**Table 2.** Odds of Reporting an LGBTQ Affirming Provider and Ever Testing for HIV among a Sample of Older HIV Positive and Negative Gay and Bisexual Men in the US South, VUSNAPS

|  | Has LGBTQ Affirming<br>Care Provider |  | Ever tested for HIV<br>(HIV Negative men only) |  |
| --- | --- | --- | --- | --- |
|  | OR | 95% CI | OR | 95% CI |
| HIV Positive | 7.102*** | [3.941-12.80] |  |  |
| Has LGBTQ Affirming Provider | -- |  | 2.260** | [1.375-3.716] |
| State of Residence |  |  |  |  |
| Georgia | 1 | [1-1] | 1 | [1-1] |
| Alabama | 0.823 | [0.477-1.420] | 0.894 | [0.432-1.846] |
| North Carolina | 1.576+ | [0.962-2.581] | 1.277 | [0.654-2.493] |
| Tennessee | 1.157 | [0.722-1.852] | 1.377 | [0.706-2.684] |
| Age | 0.986 | [0.959-1.014] | 0.969 | [0.933-1.006] |
| Education |  |  |  |  |
| High School or less | 1 | [1-1] | 1 | [1-1] |
| Some College- | 2.009 | [0.818-4.931] | 0.405 | [0.105-1.562] |
| College Degree | 2.553* | [1.060-6.149] | 1.207 | [0.310-4.698] |
| Graduate/Professional Degree | 4.620*** | [1.904-11.21] | 0.782 | [0.203-3.015] |
| Race/Ethnicity |  |  |  |  |
| White | 1 | [1-1] | 1 | [1-1] |
| Black | 0.859 | [0.390-1.892] | 1.140 | [0.311-4.178] |
| Other Person of Color | 0.585 | [0.271-1.263] | 0.430+ | [0.160-1.152] |
| Partnered | 1.716** | [1.186-2.482] | 0.683 | [0.403-1.158] |
| pseudo R-sq | 0.101 |  | 0.070 |  |
| N | 633 |  | 502 |  |

Note: Odds Ratios are exponentiated logit coefficients. + p<0.1- \* p<0.05- \*\* p<0.01- \*\*\* p<.001.

### Awareness of U=U

The most well-studied dimension of the U=U concept is sexual minority men’s *awareness* of U=U. Only about one in four (25.4%) of older gay and bisexual men in the four Southern states sampled had heard of the U=U concept; a majority (70.5%) had *<u>not</u>* heard of the U=U concept or were uncertain (4.4%). HIV positive men (56.3%) were significantly more likely to have heard of U=U compared with HIV negative men (17.5%; *χ*^*2*^= 85.829; *p*<.001). Both HIV negative and HIV positive men with an LGBTQ affirming provider were significantly more likely to have heard about U=U compared to men of the same HIV status without an affirming provider (*p*<.001; see Table 3).

**Table 3.**
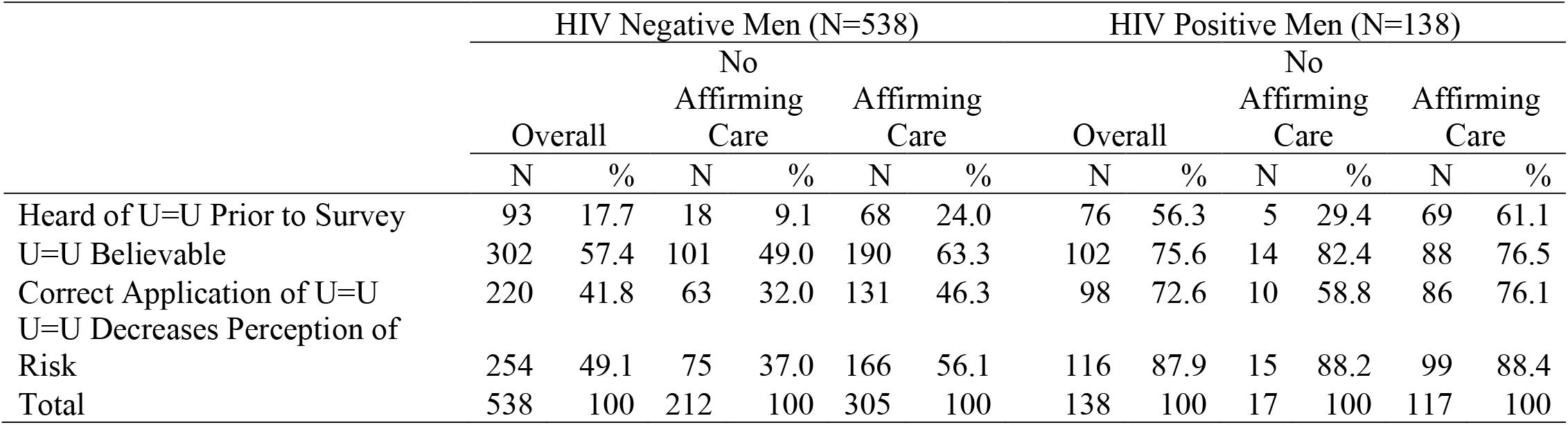
Characteristics of Older HIV Positive and HIV Negative Gay and Bisexual Men in the US South, VUSNAPS

In logistic regression analyses adjusting for other respondent characteristics and geographic variation (see Table 4), we find that HIV negative men with an affirming care provider were more than 3 times more likely to have heard of U=U (OR=3.13; 95% CI=1.75-5.61). Even among HIV positive men, who had high overall awareness of U=U, those with an affirming care provider were almost 5 times more likely to report having heard of U=U (OR=4.87; 95% CI=1.31-18.05).

**Table 4.** Estimates of U=U Awareness among a Sample of Older HIV Positive and Negative Gay and Bisexual Men in the US South, VUSNAPS

|  | Heard about U=U |  |  |  |
| --- | --- | --- | --- | --- |
|  | HIV Negative Men |  | HIV Positive Men |  |
|  | OR | 95% CI | OR | 95% CI |
| Has LGBTQ Affirming Provider | 3.131*** | [1.749-5.606] | 4.869* | [1.313-18.05] |
| Ever Tested for HIV | 1.816 | [0.839-3.931] | -- |  |
| State of Residence |  |  |  |  |
| Georgia | 1 | [1-1] | 1 | [1-1] |
| Alabama | 1.151 | [0.549-2.410] | 3.307+ | [0.876-12.49] |
| North Carolina | 0.564+ | [0.288-1.105] | 0.803 | [0.270-2.383] |
| Tennessee | 0.673 | [0.351-1.292] | 0.505 | [0.189-1.353] |
| Age | 0.982 | [0.945-1.021] | 1.010 | [0.944-1.081] |
| Education |  |  |  |  |
| High School or less | 1 | [1-1] | 1 | [1-1] |
| Some College- | 4.446 | [0.539-36.66] | 3.088 | [0.677-14.09] |
| College Degree | 2.688 | [0.331-21.82] | 0.869 | [0.197-3.838] |
| Graduate/Professional Degree | 3.711 | [0.461-29.85] | 1.549 | [0.338-7.093] |
| Race/Ethnicity |  |  |  |  |
| White | 1 | [1-1] | 1 | [1-1] |
| Black | 1.939 | [0.674-5.575] | 1.013 | [0.315-3.261] |
| Other Person of Color | 1.447 | [0.450-4.650] | 3.137 | [0.724-13.59] |
| Partnered | 0.671 | [0.400-1.125] | 3.478** | [1.427-8.472] |
| pseudo R-sq | 0.074 |  | 0.156 |  |
| N | 502 |  | 131 |  |

Note: Odds Ratios are exponentiated logit coefficients. + p<0.1- \* p<0.05- \*\* p<0.01-
\*\*\* p<.001.

Among those who had heard of U=U (N=166), the most common sources of initial information were 1) the internet (26.3%), 2) a television or print advertisement or story (23.1%), or 3) a health care provider (19.4%). Respondents also indicated that they had heard about U=U from community outreach and meetings, family or friends, and social networking or dating apps. We observe significant differences in how participants heard about U=U by whether the respondent also reports having an LGBTQ affirming healthcare provider (*χ*^*2*^=18.107; *p*<.01). Among those with an LGBTQ affirming provider, 21.2% heard about U=U from their healthcare provider compared with just 8.7% among those not reporting an LGBTQ affirming healthcare provider. Among HIV negative men, those who reported hearing about U=U from a healthcare provider *all* indicated that their provider was LGBTQ affirming. Sample size limitations prohibit us from further disaggregation or adjusted analyses of this outcome.

### Believability of U=U

Following a short description of the U=U concept, a majority of sexual minority men rated U=U as “very believable” (35.1%) or “somewhat believable” (26.5%). About a quarter (23.4%) were unsure and a nontrivial minority rated U=U as “very unbelievable” (5.8%) or somewhat unbelievable (9.3%). In bivariate analyses, individuals with an LGBTQ affirming care provider were significantly more likely than individuals without an affirming care provider to rate U=U as “somewhat” or “very believable” (67.0% versus 51.6%; *χ*^*2*^= 31.353; *p*<.001). This difference remains when bivariate analyses are restricted to just HIV negative men (63.3% vs 49.0%; *χ*^*2*^= 16.370; *p*<.001).

After controlling for other demographic and geographic factors in an adjusted logistic regression model (see Table 5), we find that HIV negative men with an LGBTQ affirming provider are one-and-a-half times more likely to rate the U=U concept as “somewhat” or “very believable” (OR=1.53; 95% CI=1.02-2.30).

**Table 5.** Estimates of U=U Belief among a Sample of Older HIV Positive and Negative Gay and Bisexual Men in the US South, VUSNAPS

|  | Believe U=U |  |  |  |
| --- | --- | --- | --- | --- |
|  | HIV Negative Men |  | HIV Positive Men |  |
|  | OR | 95% CI | OR | 95% CI |
| Has LGBTQ Affirming Provider | 1.528* | [1.016-2.299] | 0.408 | [0.090-1.858] |
| Heard of U=U Prior to Survey | 8.061*** | [3.754-17.31] | 1.533 | [0.573-4.100] |
| Ever Tested for HIV | 1.390 | [0.841-2.299] | -- |  |
| State of Residence |  |  |  |  |
| Georgia | 1 | [1-1] | 1 | [1-1] |
| Alabama | 0.684 | [0.364-1.284] | 0.238+ | [0.055-1.027] |
| North Carolina | 0.612+ | [0.357-1.049] | 0.280+ | [0.067-1.162] |
| Tennessee | 0.523* | [0.307-0.893] | 0.243* | [0.068-0.874] |
| Age | 0.978 | [0.949-1.008] | 0.997 | [0.921-1.080] |
| Education |  |  |  |  |
| High School or less | 1 | [1-1] | 1 | [1-1] |
| Some College- | 1.707 | [0.586-4.975] | 0.269 | [0.028-2.548] |
| College Degree | 1.457 | [0.519-4.088] | 0.280 | [0.030-2.627] |
| Graduate/Professional Degree | 1.691 | [0.600-4.772] | 0.522 | [0.051-5.389] |
| Race/Ethnicity |  |  |  |  |
| White | 1 | [1-1] | 1 | [1-1] |
| Black | 0.805 | [0.290-2.239] | 0.474 | [0.135-1.660] |
| Other Person of Color | 1.700 | [0.642-4.502] | 0.407 | [0.108-1.537] |
| Partnered | 0.820 | [0.540-1.246] | 0.715 | [0.280-1.825] |
| pseudo R-sq | 0.106 |  | 0.097 |  |
| N | 502 |  | 131 |  |

Note: Odds Ratios are exponentiated logit coefficients. + p<0.1- \* p<0.05- \*\* p<0.01- \*\*\* p<.001.

### Understanding of U=U

Participants were asked to apply the U=U concept to assess the likelihood that an HIV negative man would contract HIV in a hypothetical, condomless sexual encounter with an HIV positive man on treatment and undetectable. Individuals with an LGBTQ affirming provider were more likely to correctly identify that the HIV negative man had “no chance or almost no chance” of contracting the virus during the described sexual encounter (26.3% vs 9.4%; *χ*^*2*^=39.3850; *p*<.001). Although HIV positive men were more likely to correctly apply the U=U concept in this context, the gap in correct application by whether individuals had an affirming care provider was present for both HIV negative and HIV positive men.

In adjusted logistic regression analyses (see Table 6), we find that HIV negative men with an LGBTQ affirming care provider were about one-and-a-half times more likely to understand and correctly apply the U=U concept to a hypothetical scenario significant at the p<.1 level (OR=1.45; 95% CI=0.96-2.20).

**Table 6.** Estimates of Understanding of U=U among a Sample of Older HIV Positive and Negative Gay and Bisexual Men in the US South, VUSNAPS

|  | Understands U=U |  |  |  |
| --- | --- | --- | --- | --- |
|  | HIV Negative Men |  | HIV Positive Men |  |
|  | OR | 95% CI | OR | 95% CI |
| Has LGBTQ Affirming Provider | 1.454+ | [0.960-2.203] | 1.500 | [0.361-6.226] |
| Heard of U=U Prior to Survey | 3.579*** | [2.114-6.060] | 9.409*** | [3.039-29.13] |
| Ever Tested for HIV | 1.308 | [0.772-2.215] | -- |  |
| State of Residence |  |  |  |  |
| Georgia | 1 | [1-1] | 1 | [1-1] |
| Alabama | 0.693 | [0.376-1.278] | 0.240+ | [0.056-1.035] |
| North Carolina | 1.014 | [0.603-1.705] | 0.790 | [0.193-3.237] |
| Tennessee | 0.429** | [0.252-0.731] | 0.461 | [0.141-1.507] |
| Age | 0.986 | [0.956-1.016] | 1.001 | [0.918-1.093] |
| Education |  |  |  |  |
| High School or less | 1 | [1-1] | 1 | [1-1] |
| Some College- | 1.885 | [0.556-6.397] | 1.093 | [0.204-5.852] |
| College Degree | 1.716 | [0.522-5.637] | 2.474 | [0.475-12.89] |
| Graduate/Professional Degree | 2.408 | [0.733-7.909] | 2.295 | [0.400-13.17] |
| Race/Ethnicity |  |  |  |  |
| White | 1 | [1-1] | 1 | [1-1] |
| Black | 0.569 | [0.204-1.589] | 0.483 | [0.140-1.673] |
| Other Person of Color | 0.247* | [0.079-0.777] | 6.406 | [0.580-70.79] |
| Partnered | 0.882 | [0.581-1.339] | 1.044 | [0.363-3.000] |
| pseudo R-sq | 0.093 |  | 0.227 |  |
| N | 502 |  | 131 |  |

Note: Odds Ratios are exponentiated logit coefficients. + p<0.1- \* p<0.05- \*\* p<0.01- \*\*\* p<.001.

### Impact of U=U on Risk Perception

To assess the impact of U=U on perceived risk, we asked participants to rate their level of agreement with the statement “I would feel safe having sex with someone who is HIV-positive as long as they are receiving treatment and have reduced their viral load to a point where it is undetectable.” A majority (57.0%) of sexual minority men agreed or strongly agreed with this statement. HIV positive men (87.9%) were significantly more likely to agree or strongly agree compared with HIV negative men (49.1%; *χ*^*2*^= 64.421; *p*<.001). Among HIV negative men, those with an LGBTQ affirming care provider (56.1%) were significantly more likely to view having sex with someone who is HIV positive and undetectable as “safe” compared with HIV negative men who did not report an LGBTQ affirming provider (37.0%; *χ*^*2*^= 17.657; *p*<.001). After controlling for other respondent demographic characteristics and geographic location (see Table 7), we find having an LGBTQ affirming care provider increased the odds of feeling safe having sex with someone who is HIV positive and undetectable by almost two-and-a-half times (OR=2.02; 95% CI=1.33-3.05).

**Table 7.** Estimates of Impact of U=U on Risk Perception among a Sample of Older HIV Positive and Negative Gay and Bisexual Men in the US South, VUSNAPS

|  | U=U Decreases Perception of Risk |  |  |  |
| --- | --- | --- | --- | --- |
|  | HIV Negative Men |  | HIV Positive Men |  |
|  | OR | 95% CI | OR | 95% CI |
| Has LGBTQ Affirming Provider | 2.018*** | [1.334-3.052] | 0.251 | [0.028-2.270] |
| Heard of U=U Prior to Survey | 4.282*** | [2.413-7.601] | 3.932+ | [0.817-18.94] |
| Ever Tested for HIV | 1.958* | [1.146-3.347] | -- |  |
| State of Residence |  |  |  |  |
| Georgia | 1 | [1-1] | 1 | [1-1] |
| Alabama | 0.720 | [0.386-1.342] | 0.566 | [0.055-5.847] |
| North Carolina | 0.642 | [0.378-1.092] | 0.268 | [0.035-2.036] |
| Tennessee | 0.540* | [0.317-0.917] | 0.255 | [0.042-1.534] |
| Age | 0.980 | [0.951-1.010] | 1.022 | [0.904-1.156] |
| Education |  |  |  |  |
| High School or less | 1 | [1-1] | 1 | [1-1] |
| Some College- | 1.681 | [0.545-5.184] | 0.179 | [0.017-1.948] |
| College Degree | 1.575 | [0.530-4.676] | 1.020 | [0.079-13.15] |
| Graduate/Professional Degree | 1.425 | [0.478-4.254] | 1.314 | [0.090-19.28] |
| Race/Ethnicity |  |  |  |  |
| White | 1 | [1-1] | 1 | [1-1] |
| Black | 1.299 | [0.476-3.545] | 0.184* | [0.040-0.855] |
| Other Person of Color | 0.795 | [0.310-2.033] | 0.721 | [0.064-8.070] |
| Partnered | 0.726 | [0.478-1.103] | 1.652 | [0.359-7.609] |
| pseudo R-sq | 0.104 |  | 0.234 |  |
| N | 495 |  | 128 |  |

Note: Odds Ratios are exponentiated logit coefficients. + p<0.1- \* p<0.05- \*\* p<0.01- \*\*\* p<.001.

## Discussion

In this paper, we examine the relationship between having an LGBTQ affirming provider and several U=U related outcomes, including awareness, belief, understanding, and impact on risk perception. About two-thirds of sexual minority men in the study reported having an LGBTQ affirming healthcare provider as their primary or secondary provider. Unsurprisingly, HIV positive men were several times more likely to report having an LGBTQ affirming healthcare provider compared with HIV negative men.

Strikingly, the older gay and bisexual men in the US South surveyed by VUSNAPS are largely unaware of the U=U concept. This is especially true of HIV negative men in this study, only 17.5% of whom reported being aware of the U=U concept prior to the study. Awareness of U=U in this sample is substantially lower than other international and US surveys of people living with HIV and men who have sex with men, perhaps due to community-engagement biases in other convenience samples or lack of disaggregation of HIV negative from HIV positive men in some samples. VUSNAPS is a study of older LGBTQ adults aged 50 to 76 in a region that is disproportionately growing in HIV cases relative to the rest of the US [12], has fewer HIV and LGBTQ affirming providers [35], has more rural and suburban LGBTQ adults [36,37], and poorer access to healthcare overall [38]. Unlike other convenience sample studies, VUSNAPS purposefully recruited from a range of online and community venues that included but were not limited to HIV and LGBTQ community organizations, and, thus, the sample may reflect a population that is less-well connected to HIV care and information. Unlike other samples of substantially younger men who have sex with men, VUSNAPS also focuses exclusively on older sexual and gender minority populations in the US South.

On all U=U outcomes—awareness, belief, understanding, and impact on risk perception—we observe that HIV negative men with an affirming care have a greater likelihood of a positive outcome (see Figure 1). HIV negative men with an LGBTQ affirming care provider are also more than two times more likely to have ever received an HIV test compared to HIV negative men without an affirming care provider. Importantly, we also observe significant improvement in awareness of the U=U concept among HIV positive men who have an LGBTQ affirming care provider compared with HIV positive men who do not have an affirming care provider.

**Figure 1.**
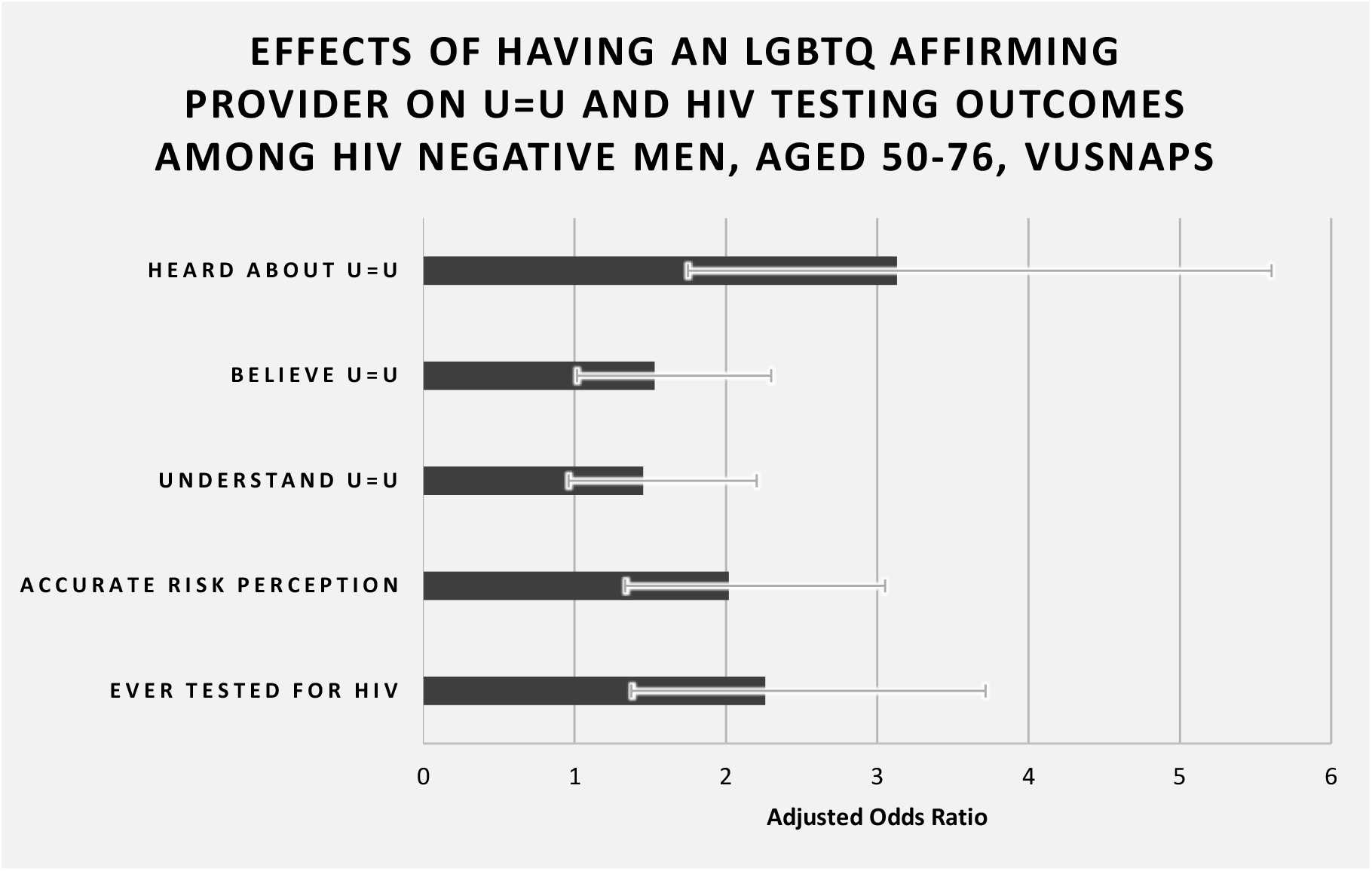
Effects of Having an LGBTQ Affirming Provider on U=U and HIV Testing Outcomes among HIV Negative Men, Aged 50-76, VUSNAPS Note: Adjusted odds ratios are presented for HIV negative men only from the analyses presented in Tables 2 and 4 to 7. Analyses control for state of residence, age, education, race/ethnicity, partner status, ever tested for HIV (except for models where this is the outcome), and ever heard of U=U (except for models where this is the outcome). All odds ratios are significant at the p<.05 level or higher except “Understand U=U”, which is significant at the p<.1 level.

There may be several mechanisms that produce these improved U=U outcomes. We find that those with an LGBTQ affirming care provider were more likely to have heard about U=U from a healthcare provider. This finding is consistent with broader findings that sexual minority patients are more likely to communicate about their specific health needs and behaviors in affirming care contexts [39]. LGBTQ affirming providers may also be more comfortable having conversations about HIV and sexual health with sexual minority men.

These findings have important implications for clinical guidance and medical education. Most physicians are comfortable treating gay patients, especially more recent medical school graduates [28,40–42]. However, additional clinical training and medical education courses on how to provide LGBTQ affirming care would likely decrease gaps in U=U awareness and understanding and may increase HIV testing among older HIV negative men in the US South. In this study, healthcare providers were among the top 3 sources of information about U=U, and men reporting an LGBTQ affirming provider were significantly more likely to have heard about U=U from their healthcare provider.

This study has some limitations. Primarily, while we see strong signals of the effects of having an LGBTQ affirming provider for HIV negative men, we lack the power to assess differences among HIV positive men, the vast majority of whom report an LGBTQ affirming provider as their primary or secondary healthcare provider. We are also unable to disaggregate experiences across race/ethnicity and sexual identity among HIV negative men. New HIV infections in Southern states are growing fastest among African American men who have sex with men. Our findings suggest but cannot confirm that greater access to LGBTQ affirming care would be particularly beneficial for increasing U=U awareness and HIV testing among this population.

Improving access to and provision of LGBTQ affirming care among sexual minority men may also reduce HIV stigma within the LGBTQ community. We find that having an LGBTQ affirming care provider increased the odds of feeling safe having sex with someone who is HIV positive and undetectable by almost two-and-a-half times (OR=2.02; 95% CI=1.33-3.05). Decreasing HIV stigma is important for the well-being of HIV positive men and increases testing among HIV negative men (cite).

## Data Availability

All data produced in the present study are available upon reasonable request to the authors or via www.vusnaps.com.

## Notes

### Competing Interest Statement

The authors have declared no competing interest.

### Funding Statement

This study was funded by the National Institute on Aging (R01AG063771).

### Author Declarations

The Vanderbilt University Human Research Protections Program gave ethical approval for this work, IRB #200117.

